# 24-hour time-use behaviors and maternal depressive symptoms during early pregnancy: Compositional data analysis

**DOI:** 10.64898/2026.10.29.26366835

**Authors:** Takashi Yoshimasu, Namhyun Kim, James Matuk, Christina M. Personette, Reagan E. Garcia, Michele D. Levine, Marquis Hawkins

**Affiliations:** Department of Epidemiology, University of Pittsburgh, Pittsburgh, PA, USA; Department of Biostatistics, Hokkaido University, Sapporo, Japan; Department of Psychology, University of Pittsburgh, Pittsburgh, PA, USA; Department of Psychiatry, University of Pittsburgh, Pittsburgh, PA, USA

**Keywords:** Compositional data analysis, physical activity, sleep, maternal depression

## Abstract

**Background:** Previous research suggested that modifiable health factors such as physical activity, sedentary behavior, and sleep (i.e., 24-hour time-use behaviors) were potential intervention targets to improve antenatal depressive symptoms. However, most previous studies examine one behavior at a time, ignoring their co-dependence. The aim of this study was to examine the association between 24-hour time-use behaviors and depressive symptoms during early pregnancy.

**Methods:** We conducted a secondary analysis using baseline data from the Health and Behaviors in Transition (HABIT) trial. Pregnant participants with pre-pregnancy body mass index ≥25 kg/m^2^ wore a wrist-worn ActiGraph for seven consecutive days to estimate moderate-to-vigorous physical activity (MVPA), light physical activity (LPA), inactivity (IA), and sleep. We used compositional data analysis with an isometric log-ratio transformation to examine associations with depressive symptoms, quantified by the Edinburgh Postnatal Depression Scale (EPDS), adjusting for maternal age, socioeconomic status, and cohabitation status. G-computation was applied to estimate the marginal mean of depressive symptoms at specified durations of each behavior, with the remaining time reallocated proportionally among the other behaviors.

**Results:** A total of 255 participants with median EPDS score of 5.0 (IQR, 2.0–8.0) had geometric mean 24-hour behaviors of MVPA = 75.5 min/day (5.2%), LPA = 311.2 min/day (21.6%), IA = 593.9 min/day (41.2%), and sleep = 459.4 min/day (31.9%). 24-hour time-use behaviors were not significantly associated with EPDS scores, and marginal mean EPDS scores showed minimal changes (MVPA 10 min: 6.51 (95% CI 4.22, 9.64) vs 40 min: 5.97 (95% CI 5.03, 6.97); sleep 300 min: 5.41 (95% CI 4.56, 6.22) vs 480 min: 5.81 (95% CI 5.23, 6.39)).

**Conclusions:** In this relatively active pregnant population, 24-hour time-use behaviors were not associated with depressive symptoms. Future research can examine later trimesters or other populations that are more sedentary or more depressed.

## INTRODUCTION

Pregnant women are particularly likely to experience poorer mental health related to elevated stress and lack of social support [1,2]. Reflecting the vulnerability of the antenatal period, antenatal depression is highly prevalent, with major depression affecting approximately 6% of U.S. individuals and minor depression affecting approximately 16% [3]. Both depression and depressive symptoms during pregnancy are associated with preterm birth, gestational diabetes, and preeclampsia [4]. Notably, depressive symptoms in early pregnancy are also among the strongest predictors of postpartum depression [5], underscoring the importance of understanding depression during this specific window. Although the etiology of antenatal depression is multifactorial, modifiable health behaviors, including poor diet, poor sleep, lack of physical activity, and smoking, contribute to approximately 32% of the variance in depressive symptoms during pregnancy [6]. Taken together, health behaviors mark as a critical factor in this particularly vulnerable period.

Physical activity, sedentary behavior (SB), and sleep are recognized as important modifiable factors for maternal health outcomes. For example, moderate-to-vigorous physical activity (MVPA) is an established factor to reduce risks for hypertensive disorders of pregnancy (HDP), gestational weight gain (GWG) that exceeds national guidelines, gestational diabetes, and postpartum depression [7,8]. However, the association with antenatal depression is currently poorly understood [7]. Previous studies suggest that higher levels of MVPA and higher sleep indicators (i.e., duration and quality) work individually as a protective factor against antenatal depression [9–12]. However, research on light physical activity (LPA) and SB is critically lacking with few studies suggesting that higher LPA is associated with lower levels of depression [13], and higher SB is associated with higher levels of depression [14]. These health behaviors are currently poorly understood with respect to their association with maternal depression.

Recently, the new paradigm—the 24-hour Activity Cycle (24-HAC) model—has emerged as a critical concept to understand an optimal 24-hour pattern of health behaviors for health outcomes [15]. While previous studies investigated the association of individual components of 24-hour time-use behaviors, two important aspects of the 24-HAC model have not been examined. First, activities are interrelated—for instance, higher levels of MVPA can reduce negative influences of SB on cardiometabolic health outcomes [16], and higher sleep quality increases effects of higher MVPA on eGWG [17], underscoring the need to incorporate other activities that are not necessarily the focus of study. Second, change in one behavior results in change in others—for example, increasing MVPA in place of SB has different implications from increasing MVPA in place of sleep [18]. Recently, compositional data analysis (CoDA) has been proposed as a methodology to examine interrelated and closed nature of these behaviors [18,19] and provide a method for examining the full range of behaviors including LPA and SB [20].

Therefore, the aim of the present study is to fill this knowledge gap by investigating the role of objectively measured 24-hour time-use behaviors—within the 24-HAC framework using CoDA—in maternal depressive symptoms during early pregnancy. While previous studies focused extensively on MVPA and sleep as individual exposures [9–12], the 24-HAC model suggests that SB and LPA, through their interrelationships with MVPA and sleep, represent an important inclusions, as changing SB and LPA may be a more readily achievable behavioral target than increasing MVPA or sleep [21]. We hypothesized that higher MVPA, LPA, or sleep, along with lower SB, will be associated with fewer depressive symptoms.

## METHODS

### Study design, settings, and participants

This was a secondary analysis using data from Optimizing Health From Pregnancy Through One Year Postpartum (HABIT; NCT03069690). The HABIT trial was a sequential multiple assignment randomized trial (SMART), designed to examine the impact of different combinations of intervention to optimize perinatal health and well-being [22]. Pregnant women in the Pittsburgh area were recruited via health clinics and online research registries. All participants (*N* = 312) completed a demographic survey, self-report measures of medical conditions, and ActiGraph data at enrollment (ActiGraph wGT3X-BT, ActiGraph L.L.C., FL, USA). Participants were then randomized to intervention for weight and self-regulation during pregnancy, or an educationally-enhanced treatment as usual. At delivery, women were re-randomized to a postpartum intervention or control.

Inclusion criteria were pre-pregnancy Body Mass Index (BMI) ≥25 kg/m² based on self-report, ≤17 weeks and 4 days of gestation, English speaker, and singleton pregnancy. Exclusion criteria were multiple gestations, pre-existing diabetes, bariatric surgery within the past three years, use of medications known to affect weight, and/or acute psychiatric symptoms necessitating immediate treatment (e.g., suicidality). This study additionally excluded participants who did not have valid days of ActiGraph data (defined as non-wearing time <8 hours per day) and on whom data on educational status, household income, or insurance status were missing. For the purposes of the current study, only data from the baseline assessment before randomization were included. Data collection for the baseline assessment occurred between February 2017 and December 2022. This study conformed to the recommendations of reports regarding compositional data analysis outlined previously [19].

Eligible participants ≥18 years old provided written informed consent; those <18 years old provided verbal assent, and a parent/legal guardian provided written informed consent. All study procedures were approved by the University of Pittsburgh Institutional Review Board (PRO16020497).

### Measurements

#### Exposure: device-based assessment of 24-hour movement behavior

MVPA, LPA, SB, and sleep were assessed using the wrist-worn ActiGraph wGT3X-BT. Participants were instructed to wear the device on the non-dominant wrist continuously at all times 24h/day for seven consecutive days. Raw triaxial acceleration measured wake-to-wake time was processed using the *GGIR* R package (version 3.3.4.) [23]. Signals were auto-calibrated to local gravity and summarized into 5-second epochs. Euclidean Norm Minus One (ENMO) was used to quantify physical activity intensity. ENMO was calculated as the Euclidean norm of the raw triaxial acceleration signals minus 1 g, with negative values truncated to zero [24]. Sleep periods were estimated using the Heuristic algorithm for Distribution of Change in Z-Angle (HDCZA) [25]. Since self-reported sleep (sleep log) was not available, we classify sleep-related behaviors [26] based on their ENMO thresholds. However, wake after sleep onset (WASO) was classified as sleep [20]. Since the wrist-worn ActiGraph is not designed to capture posture, we could not differentiate SB, which is defined as the combination of posture (i.e., sitting or lying) and activity intensity, and inactivity (IA), which is defined solely by activity intensity [27,28].

Therefore, we use IA as a proxy of SB in the present study. Physical activity intensity was classified using the ENMO thresholds which were recently validated using wrist-worn ActiGraph to classify IA, LPA, and MVPA, estimating IA as <25 mg, LPA as 25–77 mg, and MVPA as ≥78 mg [28]. Non-wear time was identified using the GGIR default algorithm. Average minutes in each movement intensity and wear time were calculated separately for weekdays and weekends. These were then weighted in a 5:2 ratio and scaled for a total of 1,440 minutes per day [29].

#### Outcome: self-report Edinburgh Postpartum Depression Scale (EPDS)

The EPDS scores were used to quantify maternal depressive symptoms. The total scores were log-transformed and used on a continuous scale. EPDS was originally developed to assess postpartum depression but later validated for antenatal depression [30]. It consists of 10 short self-report statements, and each item is scored on a four-point scale from 0 to 3, yielding a total score of 0 to 30, with higher scores indicating higher depressive symptoms [31]. The EPDS has acceptable specificity (77%) and sensitivity (85%) for predicting depression [31]. Further, this measure is well validated in pregnant populations [30].

#### Demographic information

Participants self-reported age, gestational age, racial or ethical identity, educational status, household income, insurance status, employment status, and cohabitation status. Study year was also collected. Study year was dichotomized into 2017-2019 and 2020-2022 to capture the impacts of COVID-19 on time-use behaviors and depressive symptoms [32,33]. To address multicollinearity among socioeconomic indicators, socioeconomic status (SES) was created by latent class analysis using the *poLCA* R package (version 1.6.0.2) [34], and was classified as low, middle, or high based on educational status (i.e., high school, some college, 4-year college, and graduate school), household income (i.e., ≤US$30,000, US$30,001—70,000, US$70,001— 100,000, and >US$100,000), and insurance status (i.e., provided by employers, Medicare/Medicaid, personal policy, and no health insurance).

### Statistical analysis

All statistical analyses were conducted using R (version 4.5.0). Statistical significance was based on p-values below the significance level of 0.05. Maternal baseline demographics were summarized as mean (SD) or median (Interquartile range) for continuous variables and n (%) for categorical variables.

Participant’s components were transformed into isometric log-ratio (ILR) coordinates using the *compositions* R package (version 2.0.9) [35]. The geometric mean of the four time-use components (MVPA, LPA, IA, sleep) was calculated by back transforming the arithmetic average of ILR coordinates. A linear regression model was applied for log-transformed EPDS scores as an outcome, with the ILR coordinates as predictors, adjusted for maternal age (continuous), socioeconomic status (three categories), cohabitation status (binary; living together or not), study year (binary).

We used g-computation to estimate marginal mean EPDS scores, standardized to the observed distribution of covariates (maternal age, socioeconomic status, cohabitation status, and study year). For each of the four time-use components, estimated marginal means were derived from the fitted log-linear regression model by setting that component to a counterfactual value while proportionally reallocating the remaining time among the other three components. The estimated marginal EPDS scores were then processed by a homoscedastic retransformation [36,37]. Predicted mean EPDS scores were estimated at 1-min increments across the range of each behavior, defined as the sample geometric mean ± 2 SD, restricted to the observed minimum and maximum. Point estimates were the median of 1,000 bootstrap replicates, and 95% confidence intervals were the 2.5th and 97.5th percentiles of the bootstrap distribution. The detailed method was described in the **Appendix 1**.

Compositional isotemporal substitution was used to estimate differences in EPDS scores associated with reallocating up to 60 min/day from one time-use component (e.g., IA) to another (e.g., MVPA), starting from the reference composition (i.e., the sample geometric mean) and holding the remaining components (e.g., LPA and sleep) constant. For demographic covariates, mean values were used for continuous covariates, and mode values for categorical covariates, and predicted mean EPDS scores were calculated from the fitted linear model. The difference between the two predicted values (i.e., before and after reallocation) was taken as the estimated change in EPDS associated with the reallocation. These estimates are therefore conditional on the reference covariate profile. Nonparametric bootstrapping (1,000 resamples) was used to obtain confidence intervals. Predictions were corrected by a homoscedastic retransformation.

### Sensitivity analysis

Sensitivity analysis was conducted using different inclusion criteria for valid days of ActiGraph data (≥3 weekdays and ≥1 weekend day) to examine whether the inclusion of weekend days affected the overall results [38]. The same g-computation procedure was repeated in this population.

## RESULTS

The study flow chart is presented in **Figure 1**. A total of 56 (17.9%) participants were excluded due to missingness in ActiGraph data or socio-economic indicators, resulting in 255 (81.7%) participants for the primary analysis. For the sensitivity analysis, 63 (20.2%) additional participants were excluded due to insufficient ActiGraph valid data (≥3 weekdays and ≥1 weekend day), producing 192 (61.5%) participants. Participants’ characteristics (Table 1) showed high BMI by design (Median: 31.1), and the population predominantly self-identified as Non-Hispanic White (68.2%). The baseline 24-hour time-use components indicate the sample is relatively active and sleeps within recommended levels (geometric means: MVPA = 75.5 min (5.2%), LPA = 311.2 min (21.6%), IA = 593.9 min (41.2%), and sleep = 459.4 min (31.9%)).

**Figure 1.**
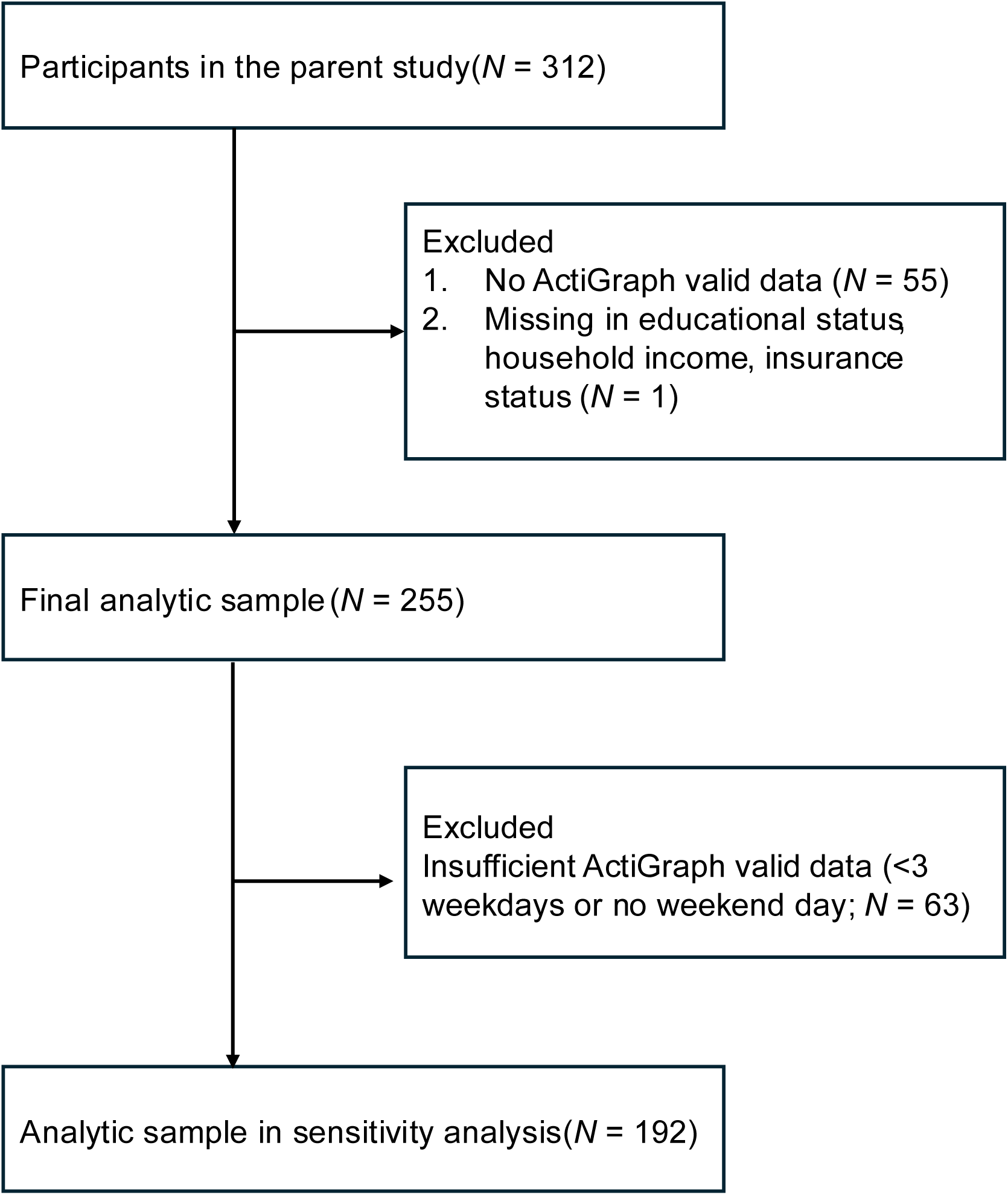
Study flow chart. Analytic sample in the primary and sensitivity analysis.

**Table 1.** Participant characteristics.

| **Characteristic** | **Statistic** | ***N* = 255** |
|---|---|---|
| **Gestational week** | Median (IQR) | 14 (11, 15) |
| **Maternal age (years)** | Mean (SD) | 30.5 (4.9) |
| **Body Mass Index (kg/m²)** | Median (IQR) | 31.1 (28.1, 36.4) |
| **Race/Ethnicity** | n (%) |  |
| Non-Hispanic White |  | 174 (68.2%) |
| Non-Hispanic Black |  | 53 (20.8%) |
| Non-Hispanic Asian |  | 1 (0.4%) |
| Hispanic |  | 14 (5.5%) |
| Other identities |  | 12 (4.7%) |
| Missing |  | 1 (0.4%) |
| **Socioeconomic status** | n (%) |  |
| Low |  | 61 (23.9%) |
| Middle |  | 109 (42.7%) |
| High |  | 85 (33.3%) |
| **Employment** | n (%) |  |
| Working |  | 196 (76.9%) |
| Not working |  | 59 (23.1%) |
| **Cohabitation status** | n (%) |  |
| Living with partner |  | 210 (82.4%) |
| Living without partner |  | 45 (17.6%) |
| **Study year** | n (%) |  |
| 2017-2019 |  | 174 (68.2%) |
| 2020-2022 |  | 81 (31.8%) |
| **24-hour time-use behaviors** |  |  |
| MVPA<sup>1</sup> (min/day) | Geometric mean (%) | 75.5 (5.2%) |
| LPA<sup>2</sup> (min/day) | Geometric mean (%) | 311.2 (21.6%) |
| IA<sup>3</sup> (min/day) | Geometric mean (%) | 593.9 (41.2%) |
| Sleep (min/day) | Geometric mean (%) | 459.4 (31.9%) |
| **EPDS<sup>4</sup> score** | Median (IQR) | 5.0 (2.0, 8.0) |

<sup>1</sup>MVPA, moderate-to-vigorous physical activity; <sup>2</sup>LPA, light physical activity; <sup>3</sup>IA, inactivity; <sup>4</sup>EPDS, Edinburgh Postnatal Depression Scale.

Likewise, the sample reported relatively few depressive symptom (median EPDS score = 5.0 (IQR, 2.0–8.0)). Demographics between those included in the primary analysis and those excluded were overall comparable (**Appendix Table 1**).

There was no statistically significant evidence that 24-hour time-use components were associated with log-transformed EPDS scores (F(3, 246) = 1.397 (*p* = 0.244)). The marginal association from g-computation is shown in **Figure 2**. If all participants took 10 minutes of MVPA while proportionally changing other behaviors, the marginal mean EPDS scores were 6.51 (95%CI: 4.21, 9.64). Likewise, 40 minutes of MVPA was associated with the marginal mean EPDS scores of 5.97 (95%CI: 5.03, 6.97). Higher LPA also suggested lower EPDS scores (240 min: 6.02 (95% CI 5.27, 6.90), 360 min: 5.47 (95% CI 4.69, 6.22)). In contrast, higher levels of IA and longer sleep suggested higher EPDS scores (IA 480 min: 5.55 (95% CI 4.85, 6.27), 660 min: 5.93 (95% CI 5.26, 6.66); sleep 300 min: 5.41 (95% CI 4.56, 6.22), 480 min: 5.81 (95% CI 5.23, 6.39)). Figure 3 shows the results of the isotemporal substitution analysis, in which 60 minutes were reallocated between each pair of the four components, relative to the geometric mean composition. Consistent with the omnibus test, individual reallocations did not indicate statistically significant associations with EPDS. Reallocating other components to MVPA or LPA suggested lower EPDS scores (60 min reallocation, IA → MVPA: EPDS -0.19 (95%CI -0.62, +0.23); IA → LPA: EPDS -0.18 (95%CI -0.48, +0.14). Reallocation between IA and sleep showed no association with EPDS scores (60 min reallocation, IA → sleep: EPDS +0.00 (95%CI -0.16, +0.22); sleep → IA: EPDS -0.01 (95%CI -0.22, +0.15) (**Appendix Table 2-3)**.

**Figure 2.**
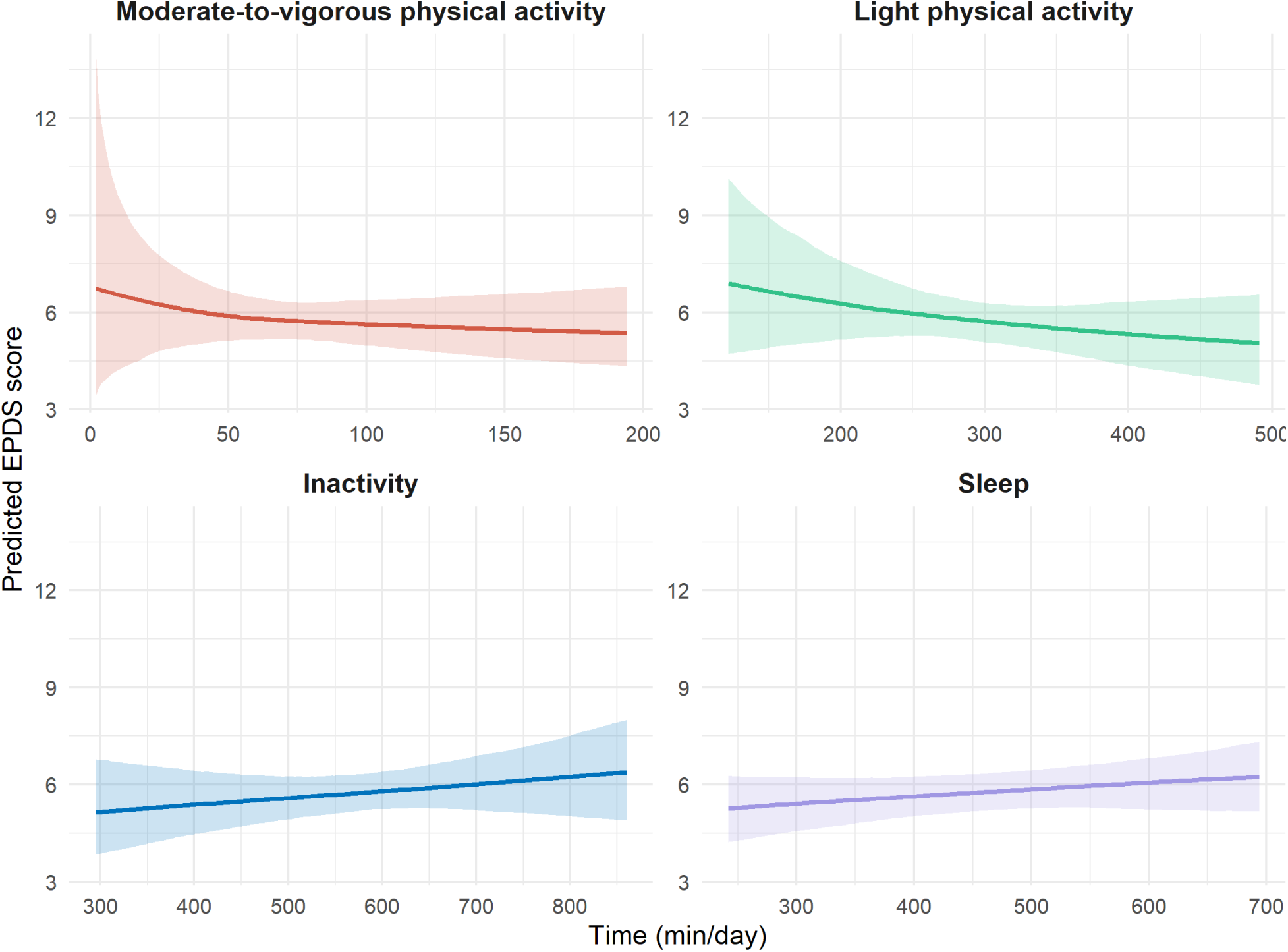
Marginal association between 24-hour time-use behaviors and maternal depressive symptoms. EPDS: Edinburgh Postnatal Depression Scale.

**Figure 3.**
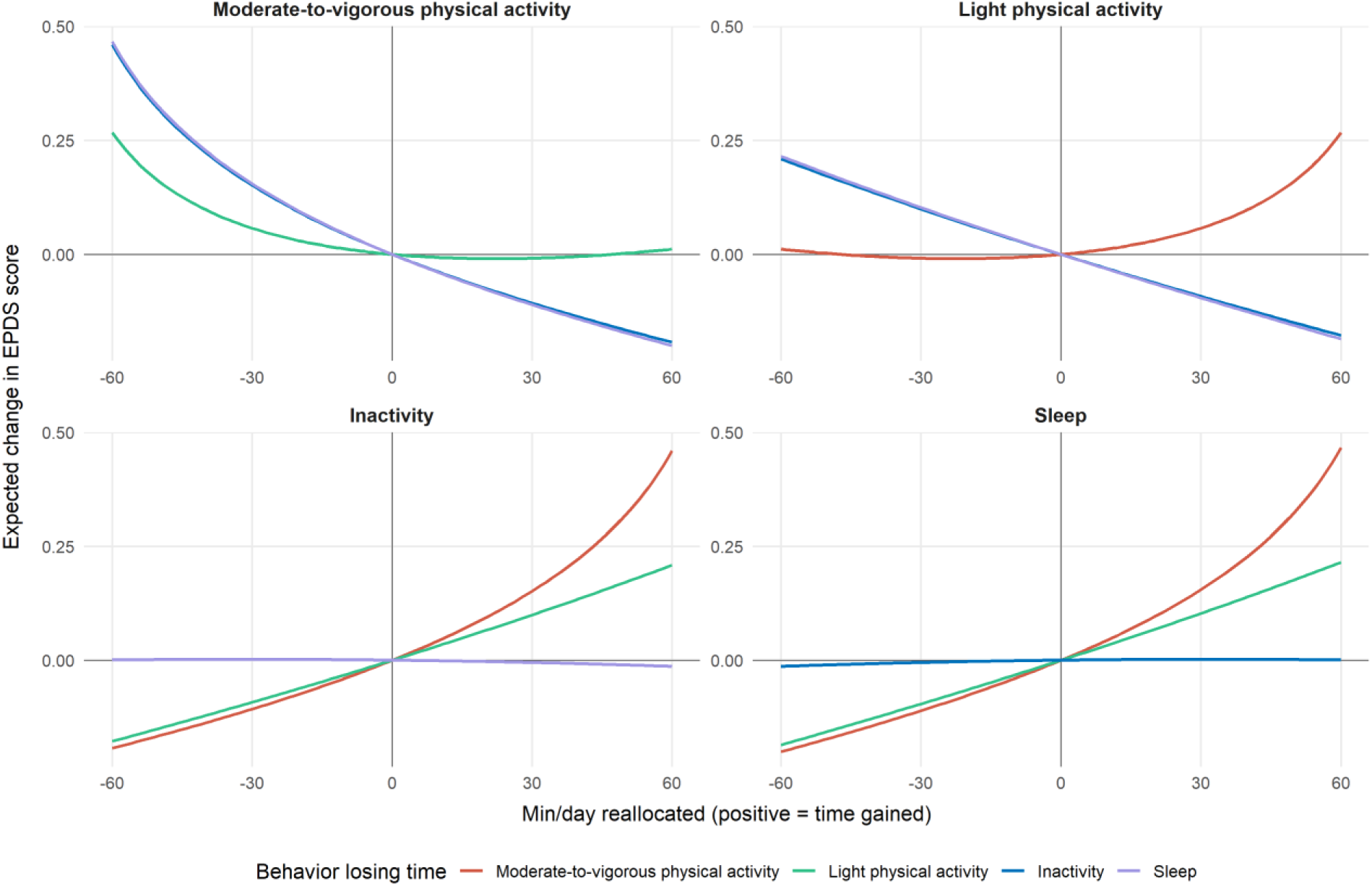
Estimated changes in EPDS scores associated with reallocating time from the geometric mean composition. EPDS: Edinburgh Postnatal Depression Scale.

Results from the sensitivity analysis are shown in **Figure 4** and the **Appendix Figure 1**. The baseline 24-hour time-use behaviors showed a geometric mean of 72.6 min (5.0%) of MVPA, 299.9 min (20.8%) of LPA, 578.5 min (40.2%) of IA, and 488.9 min (34.0%) of sleep with the median EPDS score of 5.0 (IQR, 2.0–8.0) (**Appendix Table 4**). The 24-hour time-use components were not associated with EPDS scores (F(3, 183) = 1.937 (*p* = 0.125)). Results were similar with those from primary analysis (**Figure 4**).

**Figure 4.**
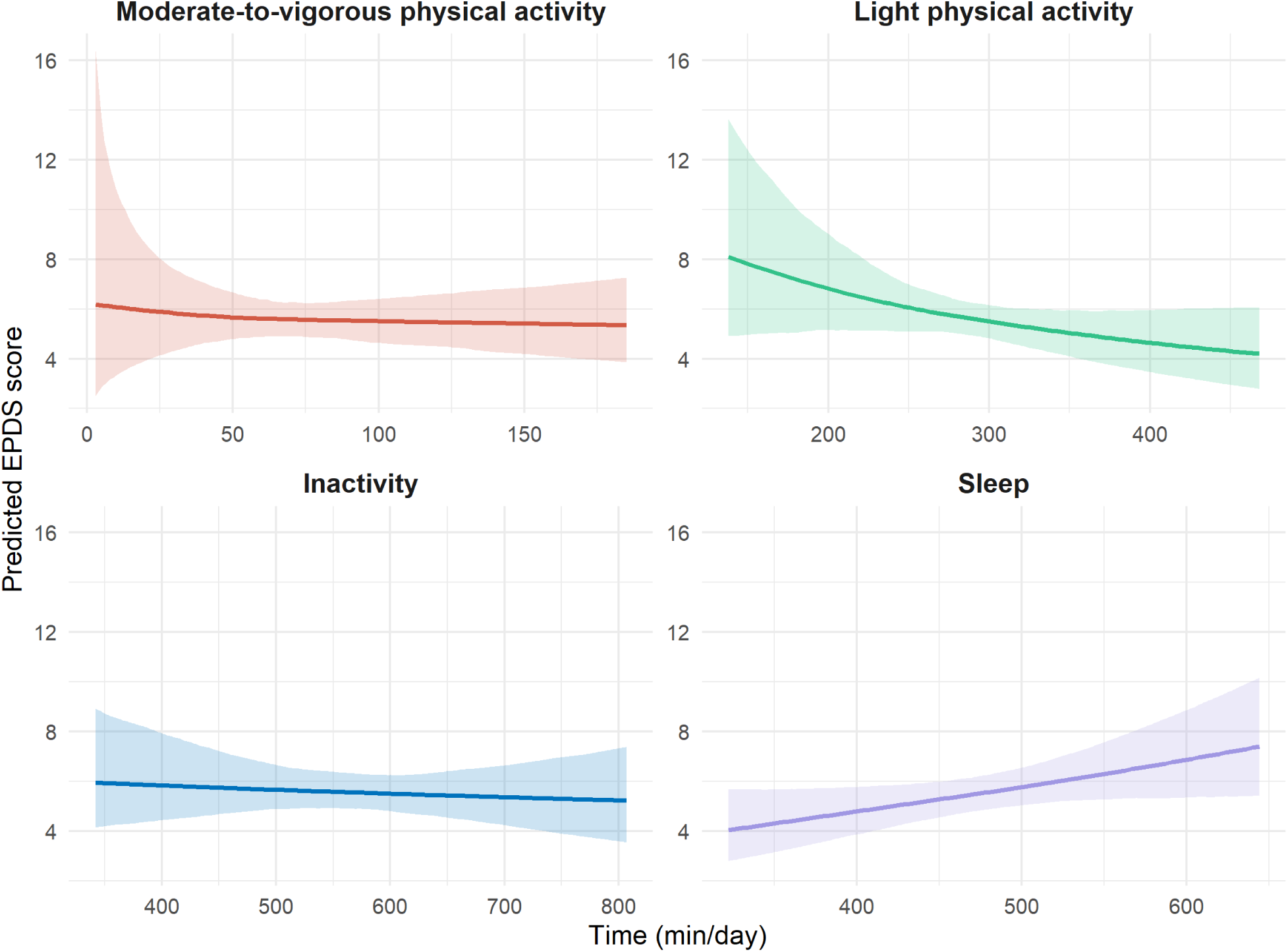
Marginal association between 24-hour time-use behaviors and maternal depressive symptoms in sensitivity analysis. EPDS: Edinburgh Postnatal Depression Scale.

## DISCUSSION

In this sample of pregnant individuals in early pregnancy, 24-hour time-use behaviors were not associated with maternal depressive symptoms. Estimated differences in EPDS scores from the g-computation and isotemporal substitution analyses, including their confidence intervals, remained well below the minimal clinically important difference (MCID) of 3–4 points [39,40]. Thus, we found no evidence of clinically meaningful associations between the reallocation of time among behaviors and depressive symptoms. This was based on device-based estimates and compositional data analysis, which accounts for the codependence of the 24-hour time-use components.

One possible explanation for the null findings is the characteristics of the study population. Most participants already met recommended levels of MVPA [41–43] and sleep [44], and depressive symptoms were generally low. In such a population, higher MVPA or longer sleep may be less likely to be associated with differences in depressive symptoms, particularly among those without clinical symptoms. The high MVPA in our sample also contrasts with previous studies of pregnant populations [20,45]. Because participants were enrolled in a randomized control trial aimed at intervening in health behaviors, they may have been motivated to stay active at enrollment. In addition, most participants were assessed during the first trimester, a period characterized by dynamic hormonal changes that are often accompanied by nausea and vomiting [46]. These pregnancy-specific factors may have obscured the associations of interest.

Measurement-related factors may also have contributed. Device-based measurement does not capture the domain of each activity. For example, walking with friends during leisure time and occupational activity can both be classified as LPA but are assumed to affect mental health differently [47]. Furthermore, because the wrist-worn ActiGraph cannot capture posture, standing, which should be classified as LPA, may have been misclassified as SB (IA). A recent study examining 24-hour time-use behaviors reported opposite associations of SB and LPA with hypertensive disorders of pregnancy [20]. If the associations of SB and LPA with depressive symptoms are also in opposite directions, such misclassification may have attenuated associations.

Although not statistically significant, the point estimates for LPA and SB (IA), which have rarely been investigated in relation to maternal depressive symptoms, were in a beneficial direction: higher LPA or lower SB (IA) was estimated to be associated with fewer depressive symptoms. This direction is consistent with previous studies highlighting the benefits of even small amounts of physical activity for antenatal depression [48,49]. However, given the small magnitude of these estimates, they should be interpreted as exploratory. In contrast, longer sleep duration tended to be associated with higher depressive symptoms, although this was also not statistically significant. Because of the cross-sectional design, this direction could reflect reverse causation, as sleep problems are more frequently reported among pregnant individuals with depression than among those without [50]. Future longitudinal studies that account for the other 24-hour time-use components are needed to examine the directionality of the relationship between sleep and depressive symptoms.

Our findings should not be interpreted as evidence against current guidelines. The Canadian 24-hour Movement Guideline acknowledged the benefits of LPA based on studies for all-cause mortality in general adult population, and recommends accumulating 150 minutes of MVPA per week, limiting SB within 8 hours, and sleeping for 7–9 hours a day [51]. The Australian guidelines also recommend staying active throughout pregnancy [52]. Because most of our participants already met these recommendations, the study was not well positioned to detect additional benefit. To our knowledge, this is the first study to use device-based measurement to examine the association between 24-hour time-use behaviors and maternal depressive symptoms in early pregnancy. LPA and SB (IA) are increasingly drawing public health attention [20,53] because they are easier to modify compared to MVPA [21,45]. Future studies with more representative and diverse pregnant populations, along with longitudinal designs, are warranted to inform time-use-based interventions to improve depressive symptoms.

## STRENGTH AND LIMITATIONS

Strengths of this study include device-based measurement of 24-hour time-use behaviors, which not only overcomes measurement bias inherent in self-report questionnaires and enables the quantification of SB (IA) and LPA, both of which are difficult to capture by self-report [15]. Additionally, CoDA accounts for the codependence of 24-hour time-use behaviors [54], and g-computation enhances the interpretability of the CoDA results by providing estimated marginal means.

Several limitations should be acknowledged. First, as described above, the population was relatively active and slept adequately, and depressive symptoms were low, which limits the generalizability of our findings and our ability to detect associations. Second, the wrist worn ActiGraph is not suited to capturing the posture, which may have led to misclassification between SB and LPA. In addition, a combination of device-based measurement and self-report is recommended to measure sleep accurately [55]. Our device-based approach could not assess sleep-related behaviors and may not have accurately captured daytime naps [26]. Third, the cross-sectional design limits causal inference because it lacks temporality, and reverse causation between time-use behaviors and depressive symptoms, which may operate bidirectionally, cannot be ruled out.

## CONCLUSIONS

In this relatively healthy sample of pregnant women, 24-hour time-use behaviors were not associated with depressive symptoms during early pregnancy. Future studies should use longitudinal designs and include populations with lower physical activity, inadequate sleep duration or quality, or higher depressive symptoms.

## Data Availability

The datasets used and/or analyzed during the current study are available from the corresponding author on reasonable request.

https://clinicaltrials.gov/study/NCT03069690

## DECLARATIONS

### Consent for publication

Not applicable.

### Authors’ contributions

Conceptualization: Takashi Yoshimasu, Namhyun Kim, and Marquis Hawkins. Methodology: Takashi Yoshimasu, Namhyun Kim, James Matuk, Reagan Garcia, and Marquis Hawkins. Formal analysis: Takashi Yoshimasu. Investigation: Michele Levine, and Marquis Hawkins. Data curation: Takashi Yoshimasu, Namhyun Kim, and Reagan Garcia.

Writing – original draft: Takashi Yoshimasu. Writing – review & editing: Namhyun Kim, James Matuk, Christina Personette, Reagan Garcia, Michele Levine, and Marquis Hawkins.

Supervision: James Matuk, Michele Levine, and Marquis Hawkins. Project administration: Michele Levine and Marquis Hawkins. Funding acquisition: Michele Levine and Marquis Hawkins.

### Acknowledgements

Takashi Yoshimasu is supported by Graduate Scholarship for Degree Seeking Students by the Japan Student Services Organization (JASSO) Student Exchange Support Program and Global Grant Scholarship by the Rotary Foundation (grant No. GG2579376). Data collection was supported by the National Institutes of Health (1R01HL132578, PI: M.D. Levine).

## List of abbreviations

BMI: Body Mass Index
IA: Inactivity
SB: Sedentary behavior
LPA: Light Physical Activity
MVPA: Moderate to Vigorous Physical Activity

## ACKNOWLEDGEMENTS

Takashi Yoshimasu is supported by Graduate Scholarship for Degree Seeking Students by the Japan Student Services Organization (JASSO) Student Exchange Support Program and Global Grant Scholarship by the Rotary Foundation (grant No. GG2579376). Data collection was supported by the National Institutes of Health (1R01HL132578, PI: M.D. Levine). Data curation was supported by Zijing Zhang.

