## Additional file 1 for "24-hour time-use behaviors and maternal depressive symptoms during early pregnancy: Compositional data analysis"

### Appendix

#### Appendix 1. Description of g-computation procedures

Let

$$\boldsymbol{X} = (X_1, X_2, X_3, X_4)^T$$

denote 24-h time-use behavior components, where

- $X_1$: Moderate-to-Vigorous Intensity Physical Activity (MVPA; min/day)
- $X_2$: Light intensity Physical Activity (LPA; min/day)
- $X_3$: Inactivity (IA; min/day)
- $X_4$: Sleep (min/day)

These components are subject to the constraint $\sum_{k=1}^{4} X_k = T$. The isometric log-ratio (ILR) transformation $\boldsymbol{Z} = ilr(\boldsymbol{X})$ is defined component-wise as

$$Z_j = \sqrt{\frac{4-j}{4-j+1}}\, ln\left(\frac{X_j}{\left(\prod_{k=j+1}^{4} X_k\right)^{1/(4-j)}}\right), j = 1, 2, 3.$$

Counterfactual exposure levels were defined as modifications of each individual's observed compositional vector $\boldsymbol{x}_i^{obs}$. Specifically, for a component $x_{ih}^{obs}(h \in \{1,2,3,4\}$; the component to be manipulated), we considered a relative change of $r_{ih}$, i.e., $a = (1 + r_{ih})\, x_{ih}^{obs}$ (a $100r\%$ change relative to the individual's observed value), while the remaining components were rescaled proportionally so that their relative composition was preserved and the total remained $T$. To satisfy the compositional constraint, a relative change by a factor of $1 + r_{ih}$ necessitates a relative change in other components by a factor of $1 - s_{ih}$, where $1 - s_{ih} = 1 - r_{ih}\frac{x_h^{obs}}{1-x_h^{obs}} = \frac{1-a}{1-x_h^{obs}}$ [1]. Therefore, a counterfactual component vector of each individual $\boldsymbol{x}_i^*$ with $x_{ih}^* = a$ is defined as

$$\boldsymbol{x}_i^* = \boldsymbol{g}_{h,a}(\boldsymbol{x}_i^{obs})$$

where

$$[\boldsymbol{g}_{h,a}(\boldsymbol{x})]_k = \begin{cases} a, & k = h, \\ x_k \dfrac{T-a}{T - x_h^{obs}}, & k \neq h. \end{cases}$$

$k$ indexes the components of the vector ($k \in \{1,2,3,4\}$)). Therefore, $\boldsymbol{z}^* = ilr\left(\boldsymbol{g}_{h,a}(\boldsymbol{x})\right)$ is the ILR representation of counterfactual values where $x_h = a$. This implies that counterfactual exposure value $\boldsymbol{z}^*$ is a function of the observed vector of time-use components and the counterfactual value of $a$.

Assuming positivity, consistency, and conditional exchangeability given covariates, our target estimand is defined as

$$E[Y^{X_h=a}] = E_{\boldsymbol{X},\boldsymbol{C}}\left[E\left[Y^{g_{h,a}(x)} \mid \boldsymbol{X} = \boldsymbol{x}, \boldsymbol{C} = \boldsymbol{c}\right]\right]$$

$$= E_{\boldsymbol{X},\boldsymbol{C}}\left[E\left[Y \mid \boldsymbol{X} = \boldsymbol{g}_{h,a}(\boldsymbol{x}), \boldsymbol{C} = \boldsymbol{c}\right]\right]$$

$$= E_{\boldsymbol{X},\boldsymbol{C}}\left[E[Y \mid \boldsymbol{Z} = \boldsymbol{z}^*, \boldsymbol{C} = \boldsymbol{c}]\right].$$

where $Y$ denotes the Edinburgh Postnatal Depression Scale (EPDS) total score, and $\boldsymbol{C}$ denotes covariates. The forth equality in the estimand definition holds since the ILR transformation is a bijective map as long as the compositional time-use vector has all strictly positive elements.

The inner expectation in the target estimand is specified through a regression model on the log scale

$$\log(Y) = \beta_0 + \boldsymbol{Z^T\beta} + \boldsymbol{C^T\gamma} + \epsilon,$$

with $\epsilon \sim (0, \sigma^2)$ and $E[exp(2\epsilon)] < \infty$. The conditional expectation on the EPDS scale is given through Duan's Smearing Method [2,3], $E[Y \mid \boldsymbol{Z} = \boldsymbol{z}_{ha}^*, \boldsymbol{C} = \boldsymbol{c}] = \exp(\beta_0 + \boldsymbol{z}_{ha}^{*\,\boldsymbol{T}}\boldsymbol{\beta} + \boldsymbol{c^T\gamma})\, E[\exp(\epsilon)]$. G-computation is used to estimate the target estimand, where predicted potential outcomes for each participant, $i$, is generated through

$$\hat{y}_i = \exp(\hat{\beta}_0 + \boldsymbol{z}_i^{*\boldsymbol{T}}\hat{\boldsymbol{\beta}} + \boldsymbol{c}_{\boldsymbol{i}}^{\boldsymbol{T}}\hat{\boldsymbol{\gamma}}) \times \frac{1}{n}\sum_{l=1}^{n}\exp(\hat{\epsilon}_l), i = 1, \dots, n,$$

where $\boldsymbol{z}_i^*$ is the ILR representation of individual's counterfactual vectors where $x_h = a$, and $\boldsymbol{c_i}$ are the observed covariate values for the participant. Finally, $\hat{E}[Y^{X_h=a}] = \frac{1}{n}\sum_{i=1}^{n}\hat{y}_i$.

**Appendix Table 1. Participant characteristics among the primary analysis and excluded**

| Characteristic | Statistic | Main analysis<br>$N = 255$ | Excluded<br>$N = 57$ |
|---|---|---|---|
| **Gestational week** | Median (IQR) | 14 (11, 15) | 15 (12, 16) |
| **Maternal age (years)** | Mean (SD) | 30.5 (4.9) | 30.3 (5.1) |
| **Body Mass Index (kg/m²)** | Median (IQR) | 31.1 (28.1, 36.4) | 31.7 (28.6, 35.0) |

| Characteristic | Statistic | Main analysis<br>*N* = 255 | Excluded<br>*N* = 57 |
|---|---|---|---|
| **Race/Ethnicity** | n (%) | | |
| Non-Hispanic White | | 174 (68.2%) | 33 (57.9%) |
| Non-Hispanic Black | | 53 (20.8%) | 16 (28.1%) |
| Non-Hispanic Asian | | 1 (0.4%) | 3 (5.3%) |
| Hispanic | | 12 (4.7%) | 3 (5.3%) |
| Other identities | | 14 (5.5%) | 2 (3.5%) |
| Missing | | 1 (0.4%) | 0 (0.0%) |
| **Educational status** | n (%) | | |
| High school | | 37 (14.5%) | 10 (17.5%) |
| Some college | | 68 (26.7%) | 16 (28.1%) |
| 4-year college graduate | | 71 (27.8%) | 15 (26.3%) |
| Graduate school | | 79 (31.0%) | 16 (28.1%) |
| **Household income** | n (%) | | |
| ≤US$30,000 | | 75 (29.4%) | 22 (38.6%) |
| US$30,001—70,000 | | 59 (23.1%) | 10 (17.5%) |
| US$70,001—100,000 | | 53 (20.8%) | 12 (21.1%) |
| >US$100,000 | | 68 (26.7%) | 12 (21.1%) |
| Missing | | 0 (0.0%) | 1 (1.8%) |
| **Insurance status** | n (%) | | |
| Provided by employer | | 164 (64.3%) | 33 (57.9%) |
| Medicare/Medicaid | | 85 (33.3%) | 22 (38.6%) |
| Personal Policy | | 5 (2.0%) | 1 (1.8%) |
| No Health Insurance | | 1 (0.4%) | 0 (0.0%) |
| Missing | | 0 (0.0%) | 1 (1.8%) |
| **Employment** | n (%) | | |
| Working | | 196 (76.9%) | 41 (71.9%) |
| Not working | | 59 (23.1%) | 16 (28.1%) |
| **Cohabitation status** | n (%) | | |
| Living with partner | | 210 (82.4%) | 48 (84.2%) |

| Characteristic | Statistic | Main analysis<br>*N* = 255 | Excluded<br>*N* = 57 |
|---|---|---|---|
| Living without partner | | 45 (17.6%) | 9 (15.8%) |
| **Study year** | n (%) | | |
| 2017-2019 | | 174 (68.2%) | 29 (50.9%) |
| 2020-2022 | | 81 (31.8%) | 28 (49.1%) |
| **EPDS[1] score** | Median (IQR) | 5.0 (2.0, 8.0) | 4.0 (2.0, 9.0) |

58 [1]EPDS, Edinburgh Postnatal Depression Scale.

59 **Appendix Table 2. The estimated change by 30 min reallocation**

| Behavior gaining time[1-3] | Behavior losing time | Estimated EPDS[3] scores (95% CI) |
|---|---|---|
| MVPA | LPA | -0.01 (-0.39, +0.38) |
| MVPA | IA | -0.11 (-0.36, +0.13) |
| MVPA | Sleep | -0.11 (-0.37, +0.14) |
| LPA | MVPA | +0.06 (-0.40, +0.63) |
| LPA | IA | -0.09 (-0.25, +0.08) |
| LPA | Sleep | -0.10 (-0.25, +0.06) |
| IA | MVPA | +0.15 (-0.20, +0.59) |
| IA | LPA | +0.10 (-0.08, +0.29) |
| IA | Sleep | -0.01 (-0.11, +0.08) |
| Sleep | MVPA | +0.16 (-0.21, +0.62) |
| Sleep | LPA | +0.10 (-0.06, +0.28) |
| Sleep | IA | +0.00 (-0.08, +0.11) |

60 [1]MVPA, moderate-to-vigorous physical activity; [2]LPA, light physical activity; [3]IA,
61 inactivity; [4]EPDS, Edinburgh Postnatal Depression Scale.

62 **Appendix Table 3. The estimated change by 60 min reallocation**

| Behavior gaining time[1-3] | Behavior losing time | Estimated EPDS[3] scores (95% CI) |
|---|---|---|
| MVPA | LPA | +0.01 (-0.66, +0.78) |
| MVPA | IA | -0.19 (-0.62, +0.23) |

| Behavior gaining time[1-3] | Behavior losing time | Estimated EPDS[3] scores (95% CI) |
|---|---|---|
| MVPA | Sleep | -0.20 (-0.64, +0.25) |
| LPA | MVPA | +0.27 (-0.94, +2.19) |
| LPA | IA | -0.18 (-0.48, +0.14) |
| LPA | Sleep | -0.19 (-0.47, +0.11) |
| IA | MVPA | +0.46 (-0.63, +2.09) |
| IA | LPA | +0.21 (-0.17, +0.62) |
| IA | Sleep | -0.01 (-0.22, +0.15) |
| Sleep | MVPA | +0.47 (-0.66, +2.18) |
| Sleep | LPA | +0.22 (-0.13, +0.61) |
| Sleep | IA | +0.00 (-0.16, +0.22) |

[1]MVPA, moderate-to-vigorous physical activity; [2]LPA, light physical activity; [3]IA, inactivity; [4]EPDS, Edinburgh Postnatal Depression Scale.

**Appendix Table 4. Maternal demographics among the primary analysis and sensitivity analysis**

| Characteristic | Statistic | Overall *N* = 255 | Sensitivity analysis *N* = 192 | Excluded *N* = 63 |
|---|---|---|---|---|
| **Gestational week** | Median (IQR) | 14 (11, 15) | 14 (10, 15) | 14 (12, 15) |
| **Maternal age (years)** | Mean (SD) | 30.5 (4.9) | 30.6 (5.0) | 30.4 (4.7) |
| **Body Mass Index (kg/m²)** | Median (IQR) | 31.1 (28.1, 36.4) | 30.7 (28.0, 35.6) | 32.7 (28.6, 37.9) |
| **Race/Ethnicity** | n (%) | | | |
| Non-Hispanic White | | 174 (68.2%) | 139 (72.4%) | 35 (55.6%) |
| Non-Hispanic Black | | 53 (20.8%) | 33 (17.2%) | 20 (31.7%) |
| Non-Hispanic Asian | | 1 (0.4%) | 0 (0.0%) | 1 (1.6%) |
| Hispanic | | 12 (4.7%) | 7 (3.6%) | 5 (7.9%) |
| Other identities | | 14 (5.5%) | 12 (6.3%) | 2 (3.2%) |
| Missing | | 1 (0.4%) | 1 (0.5%) | 0 (0.0%) |
| **Educational status** | n (%) | | | |
| High school | | 37 (14.5%) | 26 (13.5%) | 11 (17.5%) |

| Characteristic | Statistic | **Overall** *N* = 255 | **Sensitivity analysis** *N* = 192 | **Excluded** *N* = 63 |
|---|---|---|---|---|
| Some college | | 68 (26.7%) | 45 (23.4%) | 23 (36.5%) |
| 4-year college graduate | | 71 (27.8%) | 56 (29.2%) | 15 (23.8%) |
| Graduate school | | 79 (31.0%) | 65 (33.9%) | 14 (22.2%) |
| **Household income** | n (%) | | | |
| ≤US$30,000 | | 75 (29.4%) | 51 (26.6%) | 24 (38.1%) |
| US$30,001—70,000 | | 59 (23.1%) | 41 (21.4%) | 18 (28.6%) |
| US$70,001—100,000 | | 53 (20.8%) | 42 (21.9%) | 11 (17.5%) |
| >US$100,000 | | 68 (26.7%) | 58 (30.2%) | 10 (15.9%) |
| **Insurance status** | n (%) | | | |
| Provided by employer | | 164 (64.3%) | 128 (66.7%) | 36 (57.1%) |
| Medicare/Medicaid | | 85 (33.3%) | 59 (30.7%) | 26 (41.3%) |
| Personal Policy | | 5 (2.0%) | 4 (2.1%) | 1 (1.6%) |
| No Health Insurance | | 1 (0.4%) | 1 (0.5%) | 0 (0.0%) |
| **Employment** | n (%) | | | |
| Working | | 196 (76.9%) | 149 (77.6%) | 47 (74.6%) |
| Not working | | 59 (23.1%) | 43 (22.4%) | 16 (25.4%) |
| **Cohabitation status** | n (%) | | | |
| Living with partner | | 210 (82.4%) | 168 (87.5%) | 42 (66.7%) |
| Living without partner | | 45 (17.6%) | 24 (12.5%) | 21 (33.3%) |
| **Study year** | n (%) | | | |
| 2017-2019 | | 174 (68.2%) | 138 (71.9%) | 36 (57.1%) |
| 2020-2022 | | 81 (31.8%) | 54 (28.1%) | 27 (42.9%) |
| **24-hour time-use behaviors** | | | | |
| MVPA[1] (min/day) | Geometric mean (%) | 75.5 (5.2%) | 72.6 (5.0%) | 84.3 (5.9%) |
| LPA[2] (min/day) | Geometric mean (%) | 311.2 (21.6%) | 299.9 (20.8%) | 344.0 (23.9%) |
| IA[3] (min/day) | Geometric mean (%) | 593.9 (41.2%) | 578.5 (40.2%) | 636.0 (44.2%) |

| **Characteristic** | **Statistic** | **Overall** $N = 255$ | **Sensitivity analysis** $N = 192$ | **Excluded** $N = 63$ |
|---|---|---|---|---|
| Sleep (min/day) | Geometric mean (%) | 459.4 (31.9%) | 488.9 (34.0%) | 375.7 (26.1%) |
| **EPDS[4] score** | Median (IQR) | 5.0 (2.0, 8.0) | 5.0 (2.0, 8.0) | 5.0 (3.0, 8.0) |

[1]MVPA, moderate-to-vigorous physical activity; [2]LPA, light physical activity; [3]IA, inactivity; [4]EPDS, Edinburgh Postnatal Depression Scale.
